# *KIR2DL1* gene is a surrogate marker of protection against infection-related hospitalisation among HIV-1 unexposed versus exposed uninfected infants in Cameroon

**DOI:** 10.1101/2023.03.15.23287286

**Authors:** Luc-Aimé Kagoué Simeni, Clauvis Kunkeng Yengo, Rodrigue Kamga Wouambo, Janett Fischer, Oumarou M’rikam A Bessong, Joseph Fokam, Jules Clément Assob Nguedia

## Abstract

**Background:** HIV-exposed uninfected infants (HEU) experience appear more vulnerable to infections compared to their HIV-unexposed uninfected (HUU) peers, generally attributed to poor passive immunity acquired from the mother. This may be due to some genetic factors that could alter the immune system. We thus sought to determine the distribution of Killer Cells Immunoglobulin-Like Receptor (KIR) genes in HEU versus HUU, and study the association between KIR profiling and occurrence of infection-related hospitalization.

**Methods:** A cohort-study was conducted from May 2019 to April 2020 among HEU and HUU, followed-up at birth, week 6, 12, 24 and 48, in reference pediatric centers in Yaounde, Cameroon. Infant HIV status was determined, types of infections were analyzed, and 15 KIR genes were investigated using the sequence specific primer polymerase chain reaction (PCR-SSP) method. Rate of KIR genes and infection-related hospitalizations were compared in HEU versus HUU, with p<0.05 considered statistically significant.

**Results:** In this cohort, a total of 19 infection-related hospitalizations occurred in 66 infants (14.81%, 04/27 HUU and 38.46%, 15/39 HEU, p=0.037), the majority occurring during the first 24 weeks of life: 10 (25.64%) HEU and 03 (11.11%) HUU, p=0.14. At week 48 (39 HEU and 27 HUU), the relative risk (RR) for infection-related hospitalizations was 2.42 (95% CI: 1.028-5.823) for HEU versus HUU, with aOR 3.59 (95% CI: 1.037-12.448). Incidence of hospitalization was 3.2 (95% CI: 1.63–7.14) per 100 infant-months among HEU versus 1.2 (95% CI: 0.57–3.60) in HUU, and RR was 2.22 (95% CI: 0.50–9.39). *KIR2DL1* gene was significantly higher in HUU versus HEU (OR= 0.183, 95%CI: 0.053-0.629; p=0.003), and the absence of *KIR2DL1* was significantly associated with infection-related hospitalization (p<0.001; OR=0.063; 95%CI: 0.017-0.229).

**Conclusion:** Compared to HUU, the vulnerability of HEU is driving by *KIR2DL1*, indicating the protective role of this KIR against infection and hospitalizations.

## INTRODUCTION

HIV prevalence is very high in sub-Saharan Africa compared to the rest of the world. Consequently, the number of children exposed to HIV but born uninfected (HEU) is high in this region(A. L. Slogrove et al. 2016). However, studies have shown that HEU children have a 37-46% higher mortality rate than unexposed and uninfected children (Brennan et al. 2016). Prevention of mother-to-child transmission (MTCT) of the virus is one of the most important ways to control and reduce new infections in infants and children. This transmission can occur in utero during pregnancy, during delivery, or through breastfeeding (BF) (Mofenson 1997). Pediatric HIV can be as high as 45% in the absence of any intervention (Lehman et Farquhar 2007), including antiretroviral treatment, which contributes to reduce infection rate to <2% (Brocklehurst 2002). In Cameroon, the prevalence of HEU was 3.2% in 2020 (6). Several studies have shown that HEU children, although not infected with HIV, have still been affected by exposure to HIV and antiretroviral treatment. This condition is manifested in HEU children by increased clinical signs of immune dysfunction, susceptibility to multiple infections in infancy and childhood, differences in cell-mediated and humoral immunity compared to HUU children (Abu-Raya et al. 2016; Jalbert et al. 2019; Ruck et al. 2016; A. L. Slogrove et al. 2016). Natural killer (NK) cells are key players in antiviral and antitumor innate immunity. To carry out their functions, NK cells interact through killer immunoglobulin like receptors (KIR) present on their surface. These receptors are polymorphic and recognize human leukocyte antigens-1 (HLA-1) so that the interaction can produce a better immune response (Chavan et al. 2016). In humans, KIR receptors are molecules present on the NK cells surface. They are encoded by genes located on chromosome 19q13.4 of the leukocyte complex family (LRC) and clustered in a region of 150 to 200 kb. There are 17 KIR genes including two pseudogenes (*2DP1 and 3DP1*) (Wende et al. 1999). KIR receptors modulate the activity of NK cells by their activating or inhibiting function when they interact with their HLA ligand (Moretta et Moretta 2004).

KIR genes are highly polymorphic because their distribution varies from one individual to another. The same is true for genotypes and haplotypes whose repertoire varies from one individual to another and within different populations (Toneva et al. 2001; Yawata et al. 2002). Several studies have shown that exposure to antiretroviral therapy and HIV is a determinant of high mortality, morbidity and susceptibility to diseases in HEU compared to HUU (A. L. Slogrove et al. 2016; Brennan et al. 2016; Abu-Raya et al. 2016; Ruck et al. 2016; Desmonde et al. 2016; Dzanibe et al. 2019; Gabriel et al. 2019). In addition, social, parental and domestic conditions, the role of poverty, maternal and environmental risk factors have been identified by several recent reviews as explanatory factors for high mortality and morbidity among HEU children (A. Slogrove et al. 2012). But beyond these confounding factors, we may also have variations in immunity that are genetically caused (Jennes et al. 2006; Ballan et al. 2007; Boulet et al. 2008; Paximadis et al. 2011; Chavan et al. 2014). Therefore, KIR genes could be a host immunogenetic factor. The diversity of KIR genes is mainly due to the evolution and adaptation of the response to pathogens (Martinez-Borra et Khakoo 2008).

Hence this study aimed to determine the distribution of Killer Cells Immunoglobulin-Like Receptor (KIR) genes in HEU versus HUU, and study the association between KIR profiling and occurrence of infection-related hospitalizations.

## MATERIALS AND METHODS

### Study design

Within the frame of the PREVENT-IT cohort (CIPHER project, investigating the impact of *in utero* exposure to Tenofovir on the occurrence of neonatal tubulopathies in Cameroon), a cohort-study was conducted from May 2019 to April 2020 in reference pediatric centers in Yaounde, Cameroon (Cité-verte Hospital, Efoulan hospital, Biyem-Assi hospital and CASS Nkoldongo hospital). After sensitization and parental consent, HEU and HUU infants were recruited at birth at the HIV care and maternity units and follow-up at week 6, 12, 24 and 48 for clinical outcomes. Our study population was made up of two groups: HIV-1 negative infants born from HIV-1 negative mothers (Unexposed Uninfected: HUU) and HIV-1 negative infants from HIV-1 positive mother (Exposed Uninfected: HEU). Mothers of both groups of infants were tested negative for malaria, HBV and HCV infections when collecting blood. This study was approved by the Institutional Ethics Committee for Research on Human Health, of the University of Douala (Ethical clearance No. 1639IEC-UD/06/2018/T). Administrative authorization was equally obtained from the various collection sites in accordance with the ethical guidelines of the 1975 Declaration of Helsinki.

### Clinical Characteristics of Mothers and children

Clinical information from the mother-child were obtained using a structured questionnaire. This included feeding practices of the infants which can be artificial or breast, the type of delivery was documented as full-term vaginal delivery (FTVD) and caesarean section, mother on ART. The mothers were all taking first line antiretroviral treatment with the majority on tenofovir, lamivudine and effavirenz and the rest on zidovudine, lamivudine and effvirenz. Infection-associated hospitalizations, weight and height of each mother’s infant was also recorded.

All our study participants underwent blood sample collections, storage, DNA extraction, HIV screening, genotyping of KIR genes. KIR genotypes and haplotypes were determined according to the previous study Killer-cell Immunoglobulin-like Receptors (KIR) in HIV exposed infants in Cameroon (Luc-Aimé et al. 2021).

### Infection-related hospitalization

Whenever the children enrolled in our study had a clinically proven infection or illness, they reported it to us so that we could check and manage it. Therefore, each infection was noted on the child’s observation form.

### Statistical analysis

Statistical analyses was done using SPSS software (IBM SPSS, version 21.0, Chicago). Direct counting was used to determine frequencies of genes and variables. The relative risk (RR) was calculated to determine the degree of exposition to infection. Both, a cumulative incidence ratio, including only infants with complete follow-up to 12 months, as well as an incidence rate ratio, including all infants with any duration of follow-up with the denominator as number of months of follow-up, were calculated. Association between KIR genes and HIV status groups (HEU and HUU) was determined using Chi-squared or Fisher exact test as may be appropriate. Student’s t-test and Man Whitney U test as may be appropriate, were used to compare differences between means of continuous variables between HIV-unexposed uninfected and HIV-exposed uninfected groups. Association between KIR gene and infection-related hospitalization was done using Fisher exact test. Multivariate logistic regression analysis were used to determine association between KIR gene content variants and HIV status. A P value <0.05 was considered statistically significant.

## RESULTS

### Clinical and Demographic Details

66 infants (27 unexposed uninfected infants and 39 exposed uninfected infants) were enrolled and follow-up in this study. Demographic and Clinical Characteristics of both mothers and infants of the study are presented in Table 1. During study, females were overrepresented in the HEU group compared to the HUU group but not significant (56.4% vs. 55.6%, P=1.000). 92.3% of HIV-1 positive mothers with uninfected infants were on antiretroviral therapy while mother with HUU infants were not. Caesarean delivery was more represented in HEU group as compare to HUU group but not significant (10.3% vs. 7.4%, P=1.000). Breastfeeding was the most represented type of feeding. Weight was higher in HUU than HEU infants but difference was not significant (5196.29 ± 525.12 g vs. 5061.54 ± 771.74 g; P=0.433).

**Table 1:**
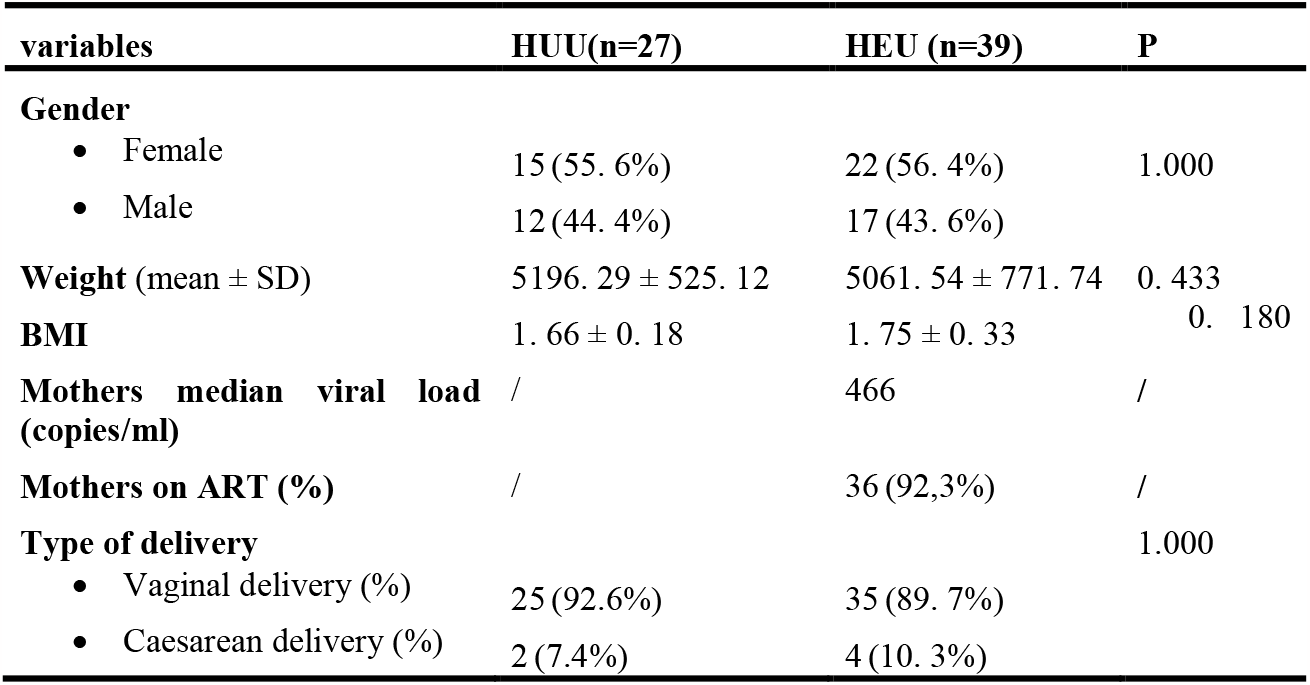
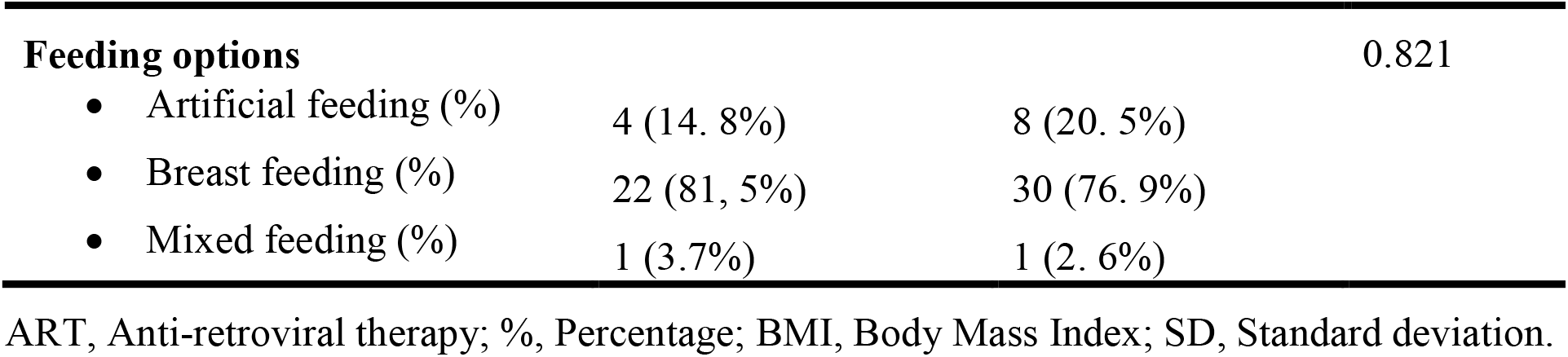
Demographic and Clinical Characteristics of both mothers and infants of the study.

| variables | HUU(n=27) | HEU (n=39) | P |
| --- | --- | --- | --- |
| **Gender** |  |  |  |
| • Female | 15 (55. 6%) | 22 (56. 4%) | 1.000 |
| • Male | 12 (44. 4%) | 17 (43. 6%) |  |
| **Weight** (mean ± SD) | 5196. 29 ± 525. 12 | 5061. 54 ± 771. 74 | 0. 433 |
| **BMI** | 1. 66 ± 0. 18 | 1. 75 ± 0. 33 | 0. 180 |
| **Mothers median viral load** / (copies/ml) | / | 466 | / |
| **Mothers on ART (%)** | / | 36 (92,3%) | / |
| **Type of delivery** |  |  | 1.000 |
| • Vaginal delivery (%) | 25 (92.6%) | 35 (89. 7%) |  |
| • Caesarean delivery (%) | 2 (7.4%) | 4 (10. 3%) |  |
| **Feeding options** |  |  | 0.821 |
| • Artificial feeding (%) | 4 (14. 8%) | 8 (20. 5%) |  |
| • Breast feeding (%) | 22 (81, 5%) | 30 (76. 9%) |  |
| • Mixed feeding (%) | 1 (3.7%) | 1 (2. 6%) |  |

ART, Anti-retroviral therapy; %, Percentage; BMI, Body Mass Index; SD, Standard deviation.

### The trend of new infections during one-year of follow-up of HUU and HEU

However, we found that 19 infection-related hospitalizations occurred in 66 infants (14.81%, 04/27 HUU and 38.46%, 15/39 HEU, p=0.037), the majority occurring during the first 24 weeks of life: 10 (25.64%) HEU and 03 (11.11%) HUU, p=0.14 (Figure 1). Lower respiratory infections accounted for 47.36% (9/19) of the hospitalizations (8 HEU and 1 HUU). Among infants who completed follow-up to 12 months (39 HEU and 27 HUU), the RR for hospitalization was 2.42 (1.028-5.823) times greater for HEU than HUU with OR 3.59 (1.037-12.448). HEU infants experienced an incidence rate of 3.2 (1.63–7.14) hospitalized infants per 100 infant-months, compared to 1.2 (0.57–3.60) in HUU infants for an RR of 2.22 (0.50–9.39).The remaining events were severe gastroenteritis (4 HEU and 1 HUU), culture confirmed urinary tract infections (2 HEU and 1 HUU), neonatal sepsis (2 HEU and 03 HUU). Two infants with gastroenteritis were coming from those on artificial feeding (Table 2).

**Figure 1:**
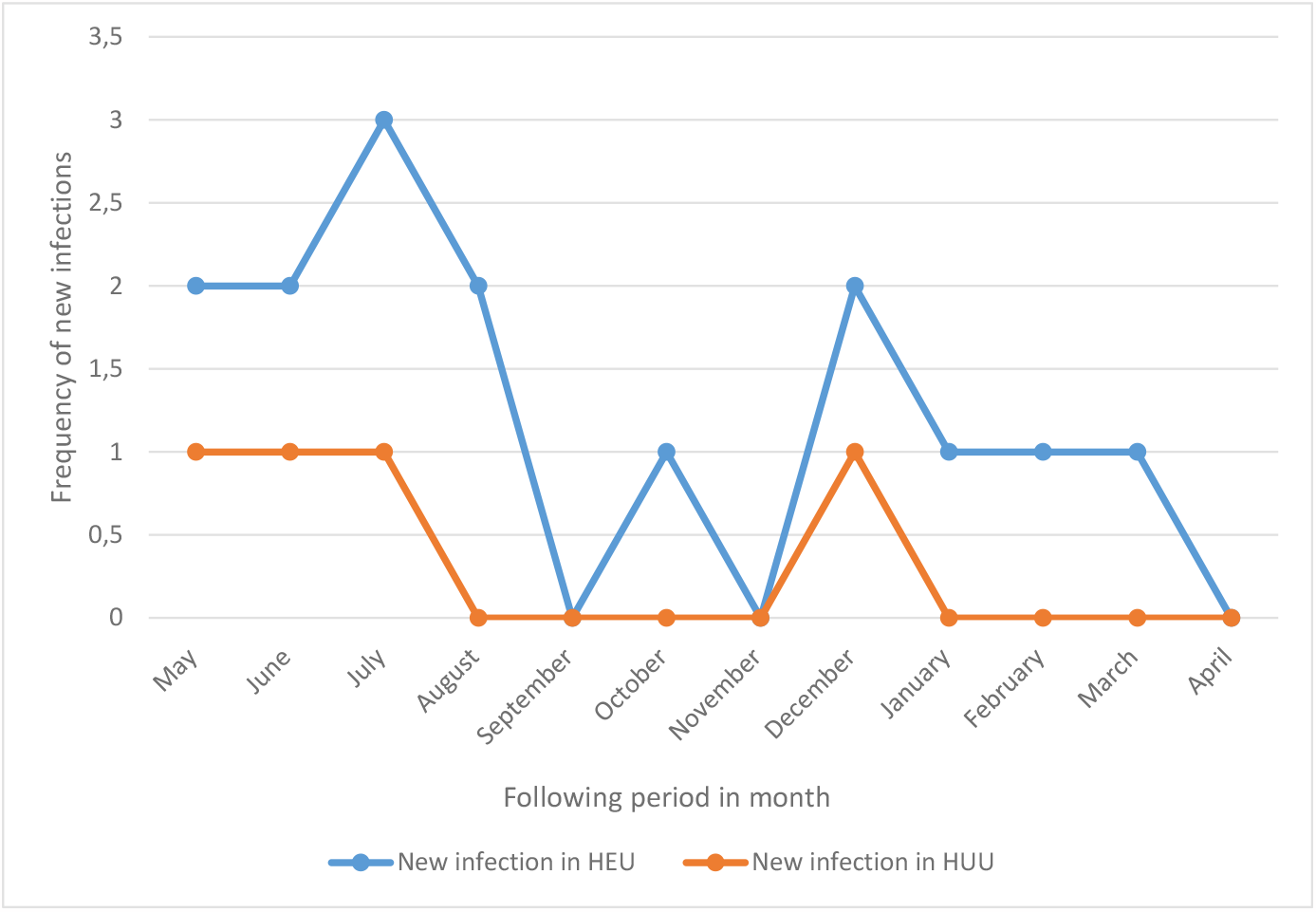
The trend of new infections during one-year of follow-up of HUU and HEU.

**Table 2:**
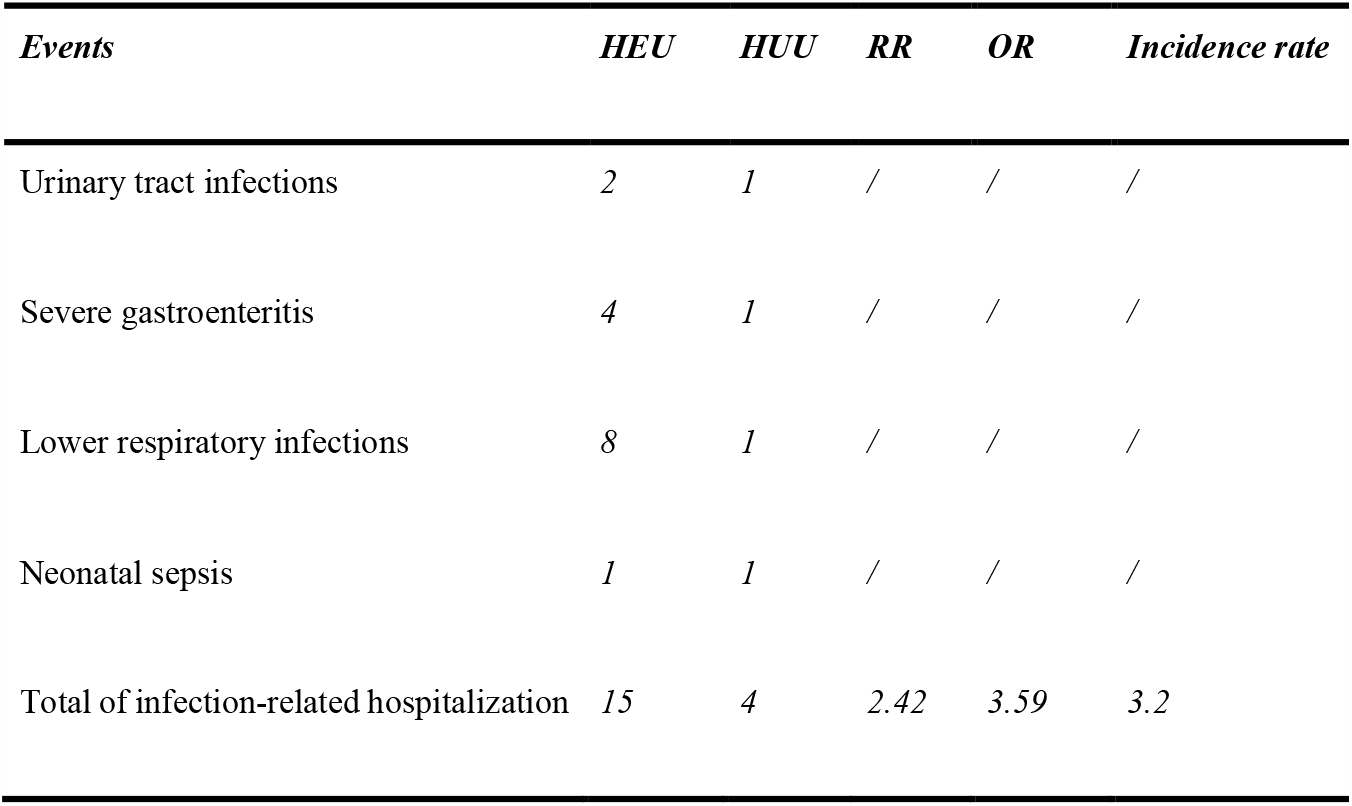
Infection-related hospitalization during 12 months of follow-up.

| *Events* | *HEU* | *HUU* | *RR* | *OR* | *Incidence rate* |
| --- | --- | --- | --- | --- | --- |
| Urinary tract infections | 2 | 1 | / | / | / |
| Severe gastroenteritis | 4 | 1 | / | / | / |
| Lower respiratory infections | 8 | 1 | / | / | / |
| Neonatal sepsis | 1 | 1 | / | / | / |
| Total of infection-related hospitalization | 15 | 4 | 2.42 | 3.59 | 3.2 |

### KIR Gene frequencies in HUU and HEU Infants

KIR gene frequencies of infants are presented in Figure 2. Frequencies of different KIR genes varied from 28.2% to 100.0%. Activating gene frequency was 28.2% to 96.3%. Frequency of inhibitory KIR genes varied from 51.3% to 96.3%. Pseudogene frequency was 64.1% to 70.4%. The frequency of the three framework genes analyzed, *KIR3DL3, KIR3DL2* and *KIR2DL4* were 100%. The KIR gene frequencies were compared between the HUU and HEU groups to determine association between presences of single KIR genes with HIV status (Figure 2). We found that the *KIR2DL1* gene was significantly more frequent in the HUU group than in the HEU (85.2% vs. 51.3%, P=0.003).

**Figure 2:**
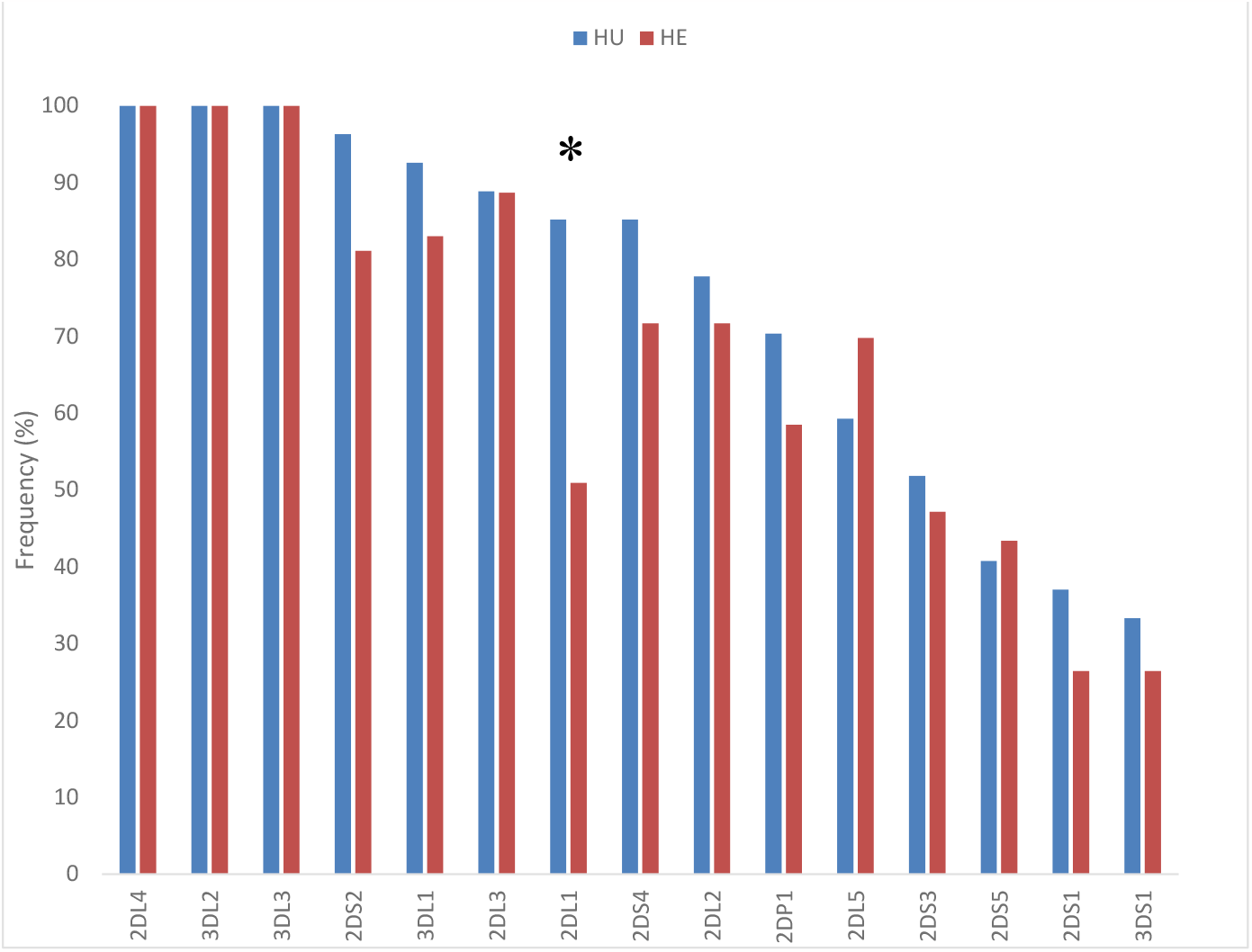
KIR gene frequencies stratified by HEU and HUU. P is the p-values comparing HIV unexposed to HIV exposed children using Chi-squared or Fisher exact test as may be appropriated

### *KIR2DL1* gene and infection-related hospitalization

The presence of *KIR2DL1* gene was more represented in people who were not hospitalized due to infections. Therefore, association between *KIR2DL1* and Infection-related Hospitalization was significant (P. value<0.001; OR=0.063; 95%CI: 0.017-0.229); the OR is closed to 0 meaning that KIR2DL1 may prevent against infection-related to hospitalization (Table 3).

**Table 3:** Relation between KIR2DL1 gene and infection-related hospitalization.

|  |  | KIR2DL1 |  | P. value | OR | 95%CI |  |
| --- | --- | --- | --- | --- | --- | --- | --- |
|  |  | No | Yes |  |  | Lower | Upper |
| Hospitalization | No | 7(14.9%) | 40(85.1%) |  |  |  |  |
|  | Yes | 14(73.7%) | 5(26.3%) |  |  |  |  |
| Total |  | 21(31.8%) | 45(68.2%) | <0.001 | 0.063 | 0.017 | 0.229 |

## DISCUSSION

Our study aimed to determine the distribution of Killer Cells Immunoglobulin-Like Receptor (KIR) genes in HEU versus HUU, and study the association between KIR profiling and occurrence of infection-related hospitalizations; HUU infants were having significantly high frequency of inhibitory gene *KIR2DL1* as compared to HEU infants. This study revealed a total of 50 genotypes. Several studies have demonstrated multiple factors that may increase the risk of infectious diseases in HEU children compared to HUU children. These factors include malnutrition, lack of breastfeeding, low lymphocyte count and high viral load, ART treatment and immune dysfunction (Dzanibe et al. 2019); Our results indicated that specific KIR genes might be associated independently with multiple diseases in HEU.

Weight mean was higher in HUU than HEU infants but difference was not significant (Table 1). This result is closed to those of Amy Slogrove, L. Afran and Ellen Moseholm, Ekali. (17, 28–30). This could be due to the effects of HIV and ART exposure on the growth outcomes of HEU children during and after pregnancy (A. L. Slogrove et al. 2016; Ekali et al. 2019). Gastroenteritis was more represented in HEU without breastfeeding contrary to respiratory infection (Table 2) showing that breastfeeding may protect against gastroenteritis. This result is close to one stipulating that, Breastfeeding is more protective against gastric infections such as diarrhea compared to respiratory infections in infancy (WHO 2000).

HUU infants were having significantly high frequencies of inhibitory gene *KIR2DL1* compared to HEU infants (Figure 1). Results confirmed that inhibitory genes are more associated with HUU status. This could be explained by the fact that activated NK cells (CD38+ CD69+) are more represented in HEU infants under 6 months of age (Haines et al. 2009). Although the inhibitory genes were more frequent at HUU status, we noticed that the majority of activating genes were represented at high frequency in HUU compared to HEU, but the differences were not significant.

Results suggest that *KIR2DL1* was more related with HUU status and even when the *KIR2DL2* and *KIR2DL3* genes were also present (Cent 3) with (Table 4). Training of NK cells by a ligand-based education model has shown that certain receptor such as KIR2DL2 can recognize HLA and suppress the subsequent expression of a second receptor *KIR2DL1*(Chang et al. 2013). Therefore, we suggest that the presence of *KIR2DL1, KIR2DL2* and *KIR2DL3* genes in NK cells decreases their inhibitory effect compared to NK cells with only *KIR2DL1* present. Thus, HUU status being associated with *KIR2DL1* reflects a reduction in NK cell inhibition.

**Table 4:**
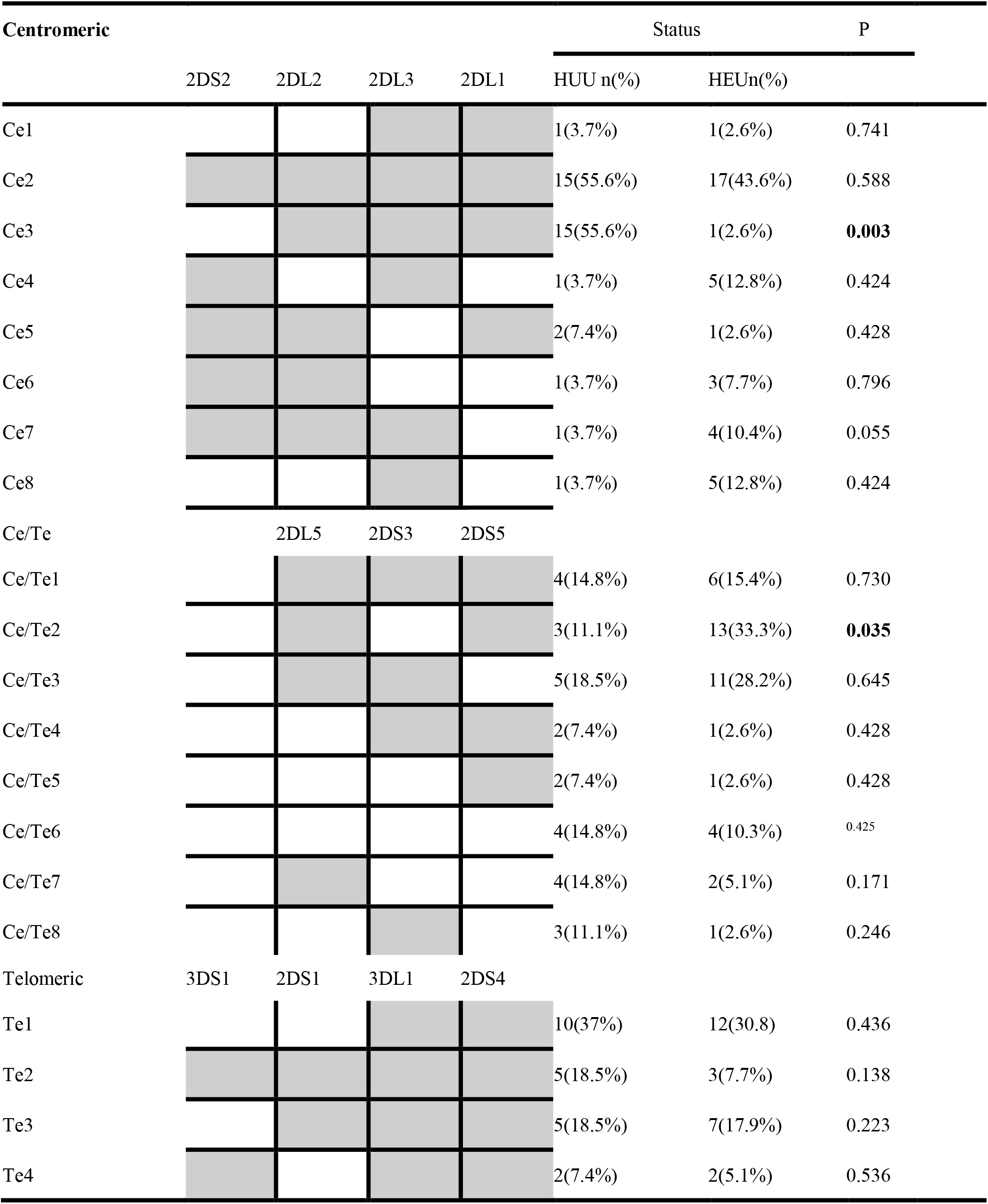
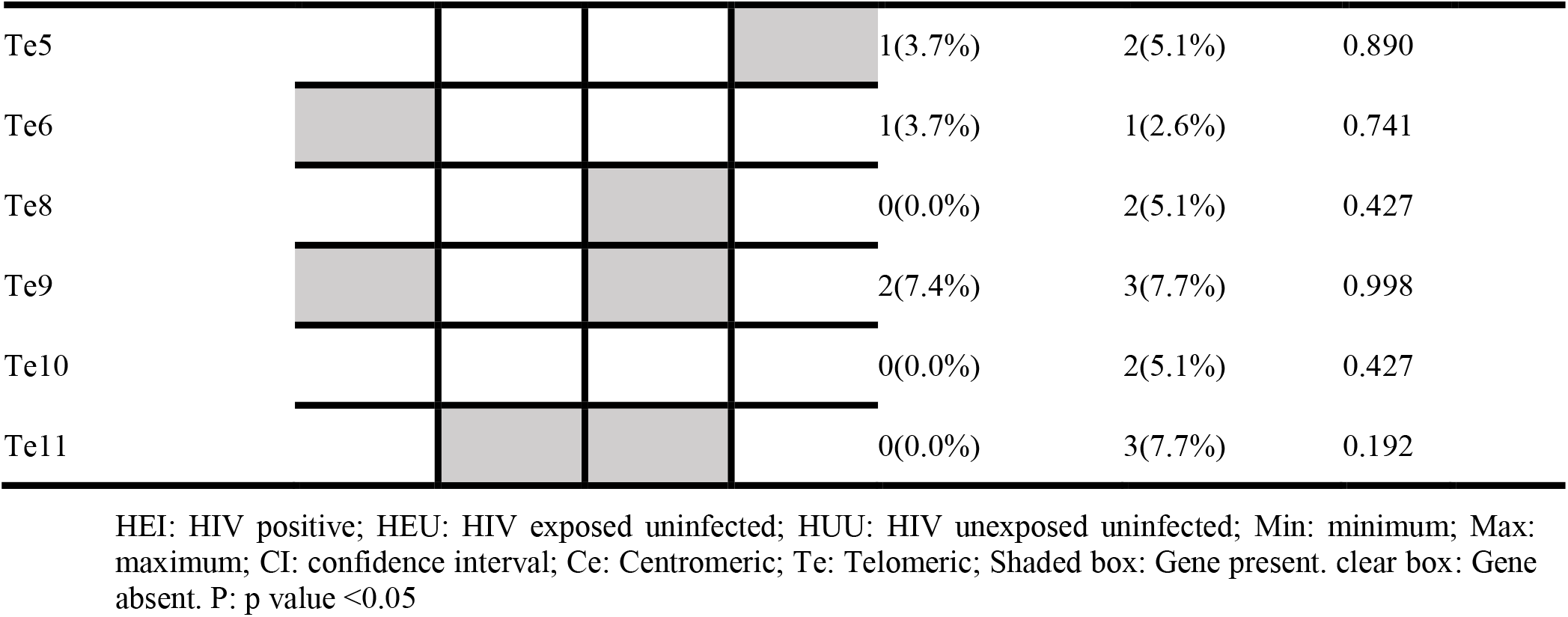
Association of KIR haplotypes locus with HIV Status.

The *KIR2DL5* gene found associated with HEU but was not sisignificant. An analysis of the telomeric/centromeric regions showed that the region including both the *2DL5* and the *KIR2DS5* genes (Ce/Te2) was also related to HEU but significantly (Table 4). Therefore we suggest that *2DS5* might have the power to increase the inhibitory effect of *2DL5*. Combination of *2DL5/2DS5* could explain the fact that NK cells are activated but still have a low cytolytic function in HEU group and being related to multiple infections.

Although there is not a statistically significant difference between the different infections depending on the status (HEU and HUU), we noted HEU infants experienced an incidence rate of 3.2 (1.63–7.14) hospitalized infants per 100 infant-months, compared to 1.2 (0.57–3.60) in HUU infants for an RR of 2.22 (0.50–9.39). At the same time, the presence of *KIR2DL1* gene was more represented in people who were not hospitalized due to infections. Therefore, *KIR2DL1* may prevent against infection-related hospitalization.

This work focused on the KIR genes trying to understand differences between HUU and HEU infants. Especially why HEU infants are susceptible to multiple infections. The strength of this study is the target population (six weeks of age) chosen before any hospitalization with regard to the results obtained. Investigating the impact of the presence and expression profiles of KIRs gene on the function of NK cells in relation to disease susceptibility as well as determine the role of NK cell in the modulation of other immune cells like dendritic cells, monocytes may be interesting leads.

## CONCLUSION

Our study which aimed to determine KIR genes and its association to multiple infections in HEU in Yaoundé-Cameroon is the first to our knowledge to be done. Compared to HUU, the vulnerability of HEU is driving by *KIR2DL1*, indicating the protective role of this KIR against infection and hospitalizations.

## Data Availability

All data produced in the present work are contained in the manuscript

## 2 Author Contributions

LA and JC designed this study. LA, CK and RK performed the search and collected data. LA and OM, re-checked data. LA and CK performed analysis. LA, wrote the first draft of the manuscript. All authors contributed to the article and approved the submitted version.

## 3 Acknowledgments

The work was supported by CIPHER Pediatric HIV Matters, Biomedsup Cameroon, and the Biotechnology Center of Yaoundé I. We would like to thank the staff of Green City, Efoulan, Mbiyem-Assi, and CASS Nkoldongo hospitals for their tremendous supports and collaborations..

